# Rasch Validation of the Warwick-Edinburgh Mental Well-being Scale (WEMWBS) in Community-Dwelling Adults and in Adults with Stroke in the US

**DOI:** 10.1101/2022.04.18.22274001

**Authors:** Wei Deng, Sydney Carpentier, Jena Blackwood, Ann Van de Winckel

**Affiliations:** Division of Rehabilitation Science, Department of Rehabilitation Medicine, Medical School, University of Minnesota, Minneapolis, Minnesota, United States of America; Division of Physical Therapy, Division of Rehabilitation Science, Department of Rehabilitation Medicine, Medical School, University of Minnesota, Minneapolis, Minnesota, United States of America

**Keywords:** Mental Well-being, Stroke, Healthy Volunteers, Community, Rasch, Validation Studies

## Abstract

**Background:** With the recent ongoing global COVID-19 pandemic and political divide in the United States (US), there is an urgent need to address the soaring mental well-being problems and to promote positive well-being. The Warwick-Edinburgh Mental Well-being Scale (WEMWBS) measures the positive aspects of mental health. Previous studies confirmed its construct validity, reliability, and unidimensionality with confirmatory factor analysis. Four studies have performed a Rasch analysis on the WEMWBS, but none of them tested adults in the US. The goals of our study are to use Rasch analysis to validate the WEMWBS in the general US population and in adults with stroke.

**Methods:** We recruited community-dwelling adults and adults with chronic stroke with upper limb hemiplegia or hemiparesis. We used the Rasch Unidimensional Measurement Model (RUMM) 2030 software to evaluate item and person fit, targeting, person separation reliability (PSR), and differential item functioning (DIF) for sample sizes of at least 200 persons in each subgroup.

**Results:** After deleting two items, the WEMBS analyzed in our 553 community-dwelling adults (average age 51.22±17.18 years; 358 women) showed an excellent PSR=0.91 as well as person and item fit, but the items are too easy for this population (person mean location=2.17±2.00). There was no DIF for sex, mental health, or practicing breathing exercises. In the 37 adults with chronic stroke (average age 58±13; 11 women) the WEMWBS had a good item and person fit, and PSR=0.92, but the items were too easy for this group as well (person mean location=3.13±2.00).

**Conclusions:** The WEMWBS had good item and person fit but the targeting is off when used in community-dwelling adults and adults with stroke in the US. Adding more difficult items might improve the targeting and capture a broader range of positive mental wellbeing in both populations. Our pilot data in adults with stroke needs to be confirmed in a larger sample size.

## Background

In recent years, the global COVID-19 pandemic has resulted in overworked health care workers, and many adults facing serious health problems, death of loved ones, and fear of losing their job.[1] Coupled with a rise in violence caused by a political divide, the United States (US) has seen a 10% increase in prevalence of adults with serious psychological distress in 2020 compared to 2018.[2] Developing positive mental well-being and resilience has therefore become critically important.

Positive mental well-being relates to feelings of happiness and life satisfaction (i.e., hedonic aspects) as well as purpose of life, full functioning of the person with a focus on realizing one’s own abilities and goals, being productive, coping with daily life stresses, and contributing to the community (i.e., eudaimonic aspects of life).[3, 4] Purpose in life or meaning plays an important role in addressing stress, trauma, and adversity.[1]

The Warwick-Edinburgh Mental Well-being Scale (WEMWBS), developed by Tennant *et al*. (2007), assesses positive mental health, covering both hedonic and eudaimonic aspects of positive well-being.[3] The internal consistency reflected by Cronbach’s α was 0.89 and 0.91, in students and adults, respectively. Confirmatory factor analysis supported the unidimensionality of the scale.[5] WEMWBS has good high test-retest reliability (*r*=0.83), good content validity, moderately high correlations with other mental health scales, and lower correlations with scales measuring overall health.[6]

Aside from these psychometric properties obtained with classical test theory (CTT), four studies have investigated the structural validity of the WEMWBS in various countries with Rasch analysis. Rasch Measurement Theory is based on a predictive model stating that a person with a higher ability on a certain trait should have a higher probability of obtaining a higher score on the scale.[6–9] The Rasch analysis ranks the item difficulty hierarchically from easy to difficult on the same logit scale as the person’s ability.[10–12] It also transforms an ordinal scale to an interval scale providing more measurement precision.[10–12]

The four studies that analyzed the WEMWBS with Rasch Measurement Theory obtained varied results in terms of targeting and number of items that remained after the Rasch analysis was completed.[6–9] Of note, the data on the scale was acquired in different countries with possibly inherent differences in culture, which could at least partially explain this variation in results. Stewart-Brown *et al*. (2009) [6] analyzed data in adults in Scotland. They obtained item fit and good targeting (person mean location −0.48±1.22). Bartram *et al*. (2013) [9] analyzed data of veterinarians in the UK and presented a short 7-item unidimensional scale that fit the model, called the Short Warwick Edinburgh Mental Well-Being Scale (SWEMWBS). However, the items were too easy for this group (i.e., person mean location 1.15±1.56). Houghton *et al*. (2015) [7] reported on a 10-item scale in adults in Western Australia with 3 misfitting items. Targeting was not reported. Finally, Wicaksono *et al*. (2021) [8] reported on the original 14-item scale with no misfitting items but the items were too easy for adults in Indonesia (i.e., person mean location 2.67±1.56). To our knowledge, there are no studies on WEMWBS data in the US population.

Aside from mental health problems in the general adult population, adults who experience a stroke are particularly vulnerable to depression, with approximately 30% of stroke survivors experiencing post-stroke depression at any given time.[13] Post-stroke depression is related to poor rehabilitation outcomes. In contrast, an increase in positive emotions over a 3-month period post-stroke is associated with an increased likelihood of functional recovery, which may lead to improved quality of life.[14, 15] Therefore, it is important to measure positive mental well-being in people with stroke and to assess whether WEMWBS would be a good measure for this population, but this has not yet been investigated.

Therefore, our first aim of this study is to assess the structural validity of the WEMWBS with Rasch in community-dwelling adults in the US. We will compare our findings with prior Rasch results in other countries. Our second aim is to perform a pilot Rasch validation on the WEMWBS in adults with chronic stroke.

## Methods

### Participants

For this cross-sectional study, we recruited participants at the Minnesota State Fair and Highland Fest and through volunteer sampling using research fliers and study postings on relevant websites. We also emailed the flier to volunteers who expressed interest in research from the Brain Body Mind Lab at the University of Minnesota. Recruitment occurred from September 27, 2017 till August 12, 2020. For both community-dwelling adults and adults with stroke, we included adults between 18-99 years of age, English speaking, and able to consent. Additionally, adults with stroke were included if they had an ischemic or hemorrhagic stroke and were medically stable. We excluded participants with stroke who had severe cognitive impairments (Mini-Mental State Exam-brief version, <13/16)[16], severe aphasia[17] or apraxia[18], or other medical conditions that would preclude participation in the study.

All community-dwelling adults gave verbal consent and were quizzed on the comprehension of the content of the consent form through the University of California, San Diego Brief Assessment of Capacity to Consent (UBACC).[19] The WEMWBS questionnaire was completed either on a tablet (at Minnesota State Fair and Highland Fest) or on their personal computer at home. All completed questionnaires were stored on the secure UMN REDCap platform. Adults with chronic stroke signed consent and completed the WEMWBS questionnaire on paper as part of a new scale development research study in the Brain Body Mind Lab. The studies were approved by the University of Minnesota’s Institutional Review Board (IRB# STUDY00005849 and STUDY00000821) and they were in accordance with the Declaration of Helsinki.

### Main outcome measures

The Warwick questionnaire covers positive aspects of mental health. All 14 items have a scoring range from “0-None of the time” to “4-All of the time”. A higher score on each item indicates a more positive attitude towards life. We collected demographic information, and whether participants currently practiced mindfulness, breathing exercises, or body awareness exercises (e.g., Yoga, Qigong, Pilates). We inquired whether they had current pain conditions or current mental health conditions.

### Statistical analysis

Following the recently accepted guidelines for reporting Rasch analyses, we report on structural validity and unidimensionality with overall fit, item and person fit, examining the presence of reversed thresholds, person separation reliability (PSR), differential item functioning (DIF), principal components analysis of residuals (PCAR), targeting, floor, and ceiling effect.[11, 12] Unidimensionality refers to the fact that all items should measure one construct. Item-trait interaction measures the overall fit of the scale to the Rasch model using Chi-square statistics. A non-significant *p*-value indicates the scale fits the model. However a large sample size can influence this *p*-value even when all items fit the model. Individual person and item fit are reported through Chi-square statistics. Residuals greater than 2.5 or smaller than 2.5 indicate item redundancy and item misfit, respectively.[10] Item fit analysis takes into account Bonferroni corrections for multiple comparisons.[20] Disordered thresholds of scoring categories can be corrected by merging adjacent categories in order to improve fit to the model.[10, 20] PSR evaluates how well individuals or groups of different ability levels can be distinguished from each other.[21] DIF occurs when the hierarchies of items are significantly different between two sample subgroups (e.g., men versus women) for sample sizes of at least 200 persons in each subgroup. DIF is calculated with an analysis of variance (ANOVA) with Bonferroni correction.[20] Further evidence of unidimensionality can be evaluated with the Principal Component Analysis of Residuals (PCAR), which refers to the extent to which covariance in the residuals is random and not explained underlying constructs than the one that is being measured.[10, 22] In that case, the expected eigenvalue is less than 2, and the percent variance explained by the first component is less than 10%. If those criteria are not met, then dependent t-tests between the 2 subsets of items with positive and negative loadings on the first residual component are performed. We would confirm unidimensionality if less than 5% of these tests are significant. A scale is well-targeted when the person mean location is between −0.5 and 0.5 logits, and thus matching the average difficulty of the items.[23] Floor and ceiling effects need to be reported when at least 15% of the sample obtains a minimum or maximum score of the scale.[24] We used Rasch Unidimensional Measurement Model (RUMM) 2030 software (RUMM Laboratory, Perth, WA, Australia) for all Rasch Measurement Theory analyses.

## Results

We recruited 553 community-dwelling adults and 37 adults with stroke. The characteristics of the demographic and clinical information of all participants is presented in ***Table 1***.

**Table 1.** Demographic and clinical characteristic of participants by group.

|  | **Community-dwelling adults**<br>**(n=533)** | **Adults with**<br>**chronic stroke**<br>**(n=37)** |
| --- | --- | --- |
| **Age (years, Mean±SD)** | 51.22±17.18 | 57.97±12.97 |
| **Years post-stroke (Mean±SD)** | N/A | 3.76±2.82 |
| **Ischemic/hemorrhagic stroke (n)** | N/A | 29/8 |
| **Sex (n)** |  |  |
| Male | 194 | 26 |
| Female | 358 | 11 |
| Other | 1 | 0 |
| **Ethnicity (n)** |  |  |
| Hispanic or Latino | 8 | 0 |
| Not Hispanic or Latino | 545 | 37 |
| **Racial background (n)** |  |  |
| American Indian or Alaska Native | 2 | 0 |
| Asian | 30 | 2 |
| Black/ African American | 11 | 1 |
| Hawaiian or other Pacific Islander | 0 | 0 |
| White | 491 | 31 |
| Multi-racial | 10 | 2 |
| Other | 9 | 1 |
| **Pain (n)** | 113 | 19 |
| **Mental health conditions (n)** | 230 | 3 |
| **Current breathing exercise (n)** | 225 | 15 |
| **Current mindfulness exercise (n)** | 181 | N/A |
| **Current body awareness training (n)** | 180 | N/A |

*Legend: N/A = not assessed*

### Rasch Measurement Theory

The iteration analysis displays the step-by-step approach taken for the Rasch analysis ***(Additional file 1)***. The main results are described below.

For our first analysis in community-dwelling Americans, none of the 14 items displayed disordered thresholds. Two items were misfitting: item 1 “*I have been feeling optimistic about the future*” and item 5 “*I have had energy to spare*.” After deleting items 1 and 5, all items fit the model and only 2.71% of persons were misfitting. The hierarchy of the item difficulty is presented in ***Figure 1***, with the easiest items starting at the top and the hardest items at the bottom. The item logit location and fit statistics are presented in ***Table 2***. There was no floor or ceiling effect, but the person mean location ± standard deviation was 2.17±2.00 logits, meaning that the items were too easy for this population ***(Figure 2)***. The PSR was 0.91, indicating that we can distinguish individuals with different positive mental health levels. PCAR’s eigenvalue was 2.04 with 16.97% variance explained by the first component. The paired t-test revealed that 7.59% of the persons had significantly different logit locations on the two subtests. These results presume the existence of two dimensions in the scale. We calculated DIF for sex (men; women), mental health conditions (yes; no), and current practice of breathing exercises (yes; no) because they all had subgroup samples sizes of at least 200 persons each. No DIF was found.

**Table 2.** Item fit statistics of the WEMWBS in community-dwelling adults in the US.

| **Item**<br>**number** | **Item descriptions** | **Item**<br>**location**<br>**(logits)** | **SE** | **Fit**<br>**Residuals** | ***p*-value** |
| --- | --- | --- | --- | --- | --- |
| Item 2 | I've been feeling useful | 0.02 | 0.07 | 1.90 | 0.17 |
| Item 3 | I've been feeling relaxed | 0.93 | 0.07 | 2.72 | 0.01 |
| Item 4 | I've been feeling interested<br>in other people | -0.03 | 0.07 | 1.24 | 0.21 |
| Item 6 | I've been dealing with<br>problems well | -0.18 | 0.08 | -0.13 | 0.84 |
| Item 7 | I've been thinking clearly | -0.18 | 0.08 | -0.37 | 0.84 |
| Item 8 | I've been feeling good about<br>myself | -0.11 | 0.07 | -4.92 | 0.003 |
| Item 9 | I've been feeling close to<br>other people | 0.03 | 0.07 | -0.83 | 0.71 |
| Item 10 | I've been feeling confident | 0.35 | 0.07 | -5.47 | 0.002 |
| Item 11 | I've been able to make up<br>my own mind about things | -0.56 | 0.07 | 2.13 | 0.05 |
| Item 12 | I've been feeling loved | -0.12 | 0.07 | 1.18 | 0.04 |
| Item 13 | I've been interested in new<br>things | -0.25 | 0.07 | -0.84 | 0.21 |
| Item 14 | I've been feeling cheerful | 0.08 | 0.07 | -4.65 | 0.04 |

**Legend:** SE = Standard Error

**Fig. 1.**
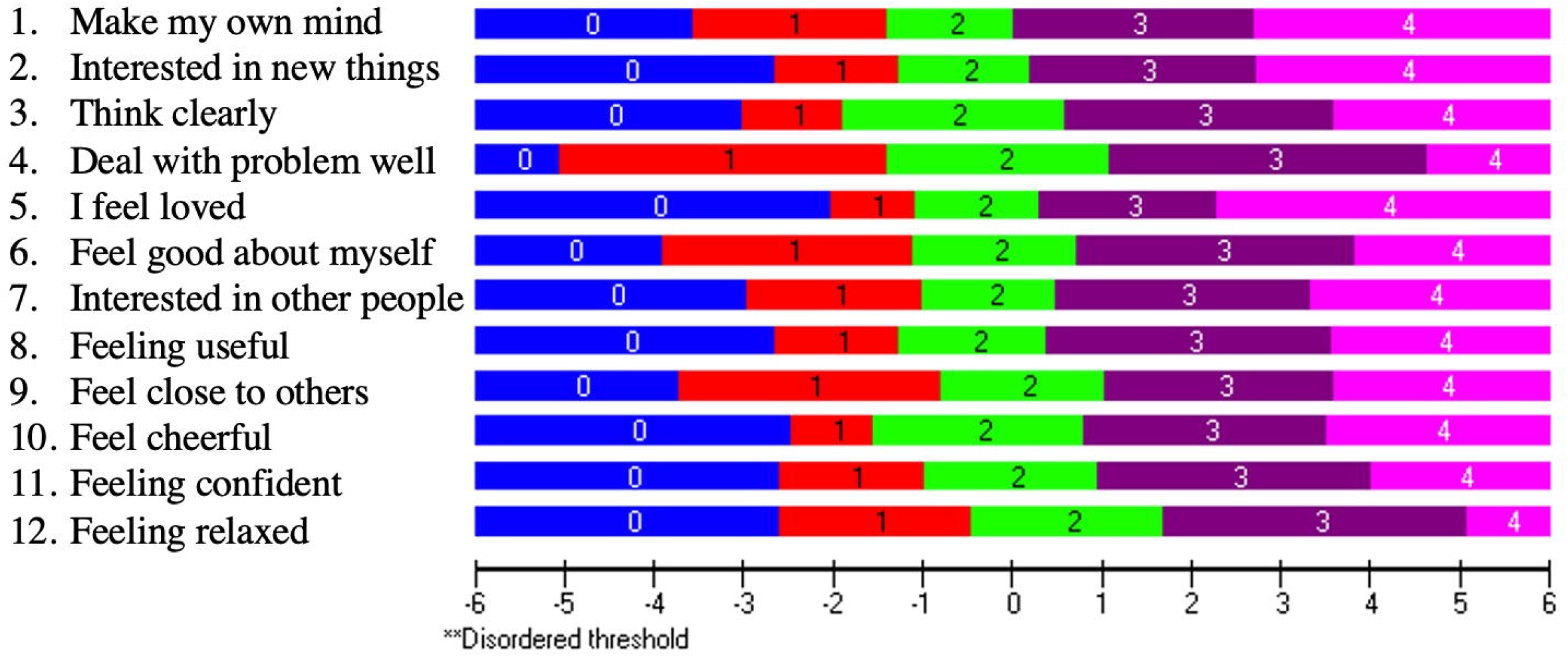
Item threshold map in community-dwelling adults in the US. The item threshold map shows the hierarchy of the item difficulty levels, with the easiest item on top (item 11 “*I’ve been able to make up my own mind about things*”) and the hardest item at the bottom (item 3 “*I’ve been feeling relaxed*”). The horizontal logit ruler demonstrates the person’s ability level of their positive mental health.

**Fig. 2.**
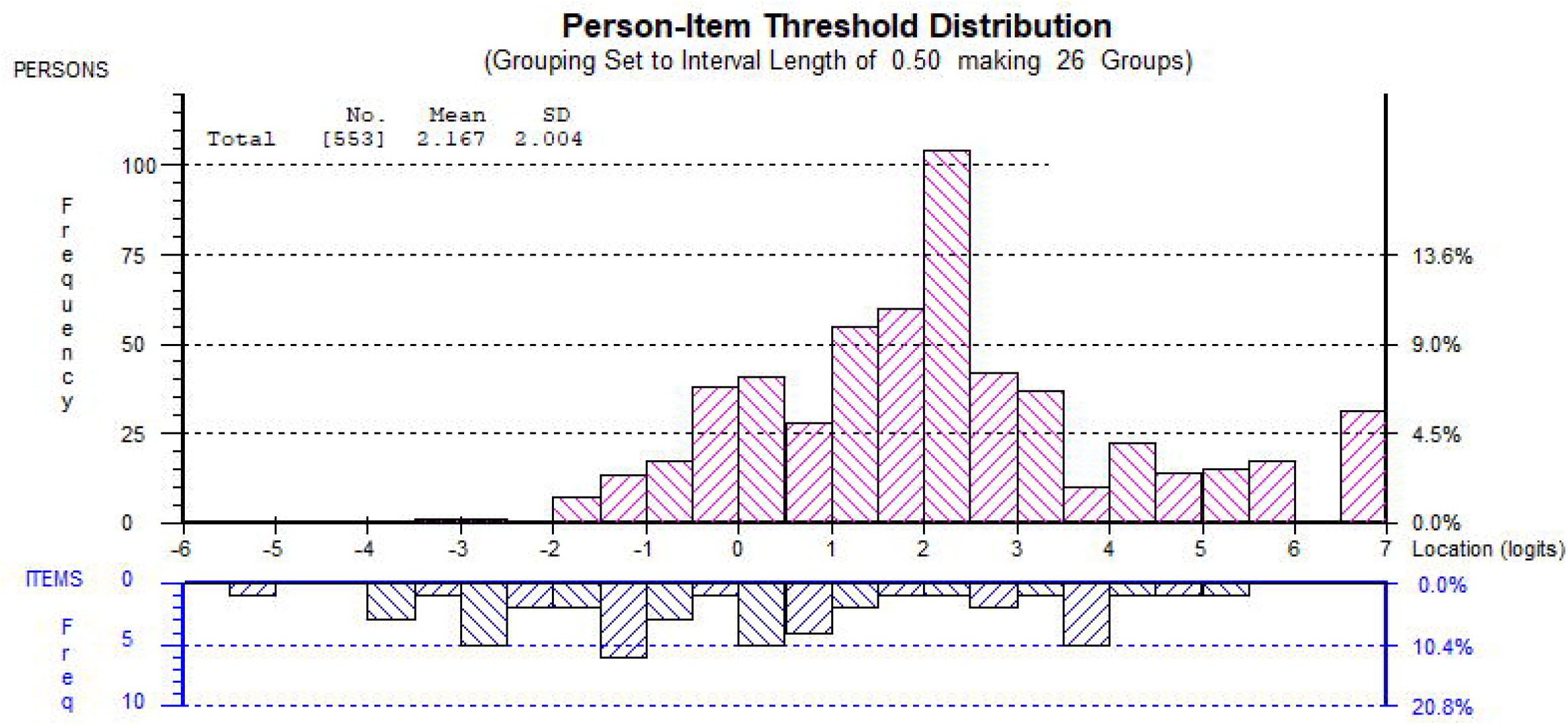
Person-item threshold distribution in community-dwelling adults in the US. The horizontal logit ruler represents both item difficulty and person ability. The pink histograms show the frequencies of the person’s ability level in terms of positive mental well-being. A higher logit value indicates the person has a higher level of positive mental well-being. The blue histograms represent the frequencies of item difficulty level, and the items are organized from the easiest on the left to the hardest on the right.

We also tested if the fit and unidimensionality would improve if we deleted items to match the 7-item SWEMWBS mentioned in previous studies. There were no misfitting items. The PCAR’s eigenvalue was 1.86 with 26.53% variance explained by the first component. The paired t-test revealed that 8.50% of the person logit pairs had significantly different locations. Additionally, the PSR dropped from 0.92 to 0.82, which would only allow researchers and clinicians to make group decisions, rather than individual decision-making.[25, 26] Moreover, the items were still too easy (person mean location 1.88±1.71). We therefore do not recommend using the 7-items scale for clinical use. We recommend that the targeting first be solved before it can be used in the clinic or for research and therefore, we do not provide a revised scoring sheet or score-to-measure table for the 12-item revised scale.

Our pilot Rasch Measurement Theory analysis in adults with chronic stroke (n=37) revealed that item 7 had reversed thresholds. After rescoring item 7 to scoring categories [00123], all items fit (***Table 3)*** and there were no more reversed thresholds ***(Figure 3)***. Only 2.70% of the people were misfitting. There was no floor or ceiling effect, and the WEMWBS had an excellent PSR of 0.92. However, the person mean location was 3.13±2.00 ***(Figure 4)***. Even though these results need to be validated in a larger study, they seem to indicate that those items are also too easy for adults with chronic stroke and very few participants choose the lowest category of “*None of the time*” or “*Rarely*” ***(Table 4)***. The PCAR analysis revealed an eigenvalue of 2.61 with 18.65% percent variance explained by the first component. The paired t-test resulted in 16.22% of pairs that had significantly different person logit locations on the 2 subtests.

**Table 3.** Item fit statistics of the WEMWBS in adults with chronic stroke.

| Item<br>number | Item descriptions | Item<br>location<br>(logits) | SE | Fit<br>Residuals | *p*-value |
| --- | --- | --- | --- | --- | --- |
| Item 1 | I've been feeling optimistic<br>about the future | -0.35 | 0.27 | -0.12 | 0.96 |
| Item 2 | I've been feeling useful | 0.53 | 0.27 | 0.62 | 0.66 |
| Item 3 | I've been feeling relaxed | 0.09 | 0.28 | 0.57 | 0.97 |
| Item 4 | I've been feeling interested<br>in other people | -0.11 | 0.26 | 0.69 | 0.54 |
| Item 5 | I've had energy to spare | 3.24 | 0.27 | 1.02 | 0.20 |
| Item 6 | I've been dealing with<br>problems well | -0.27 | 0.30 | 0.30 | 0.81 |
| Item 7 | I've been thinking clearly | -0.71 | 0.30 | 2.02 | 0.001 |
| Item 8 | I've been feeling good about<br>myself | -0.21 | 0.29 | -0.09 | 0.82 |
| Item 9 | I've been feeling close to<br>other people | 0.005 | 0.29 | 0.34 | 0.79 |
| Item 10 | I've been feeling confident | -0.11 | 0.28 | -0.78 | 0.51 |
| Item 11 | I've been able to make up<br>my own mind about things | -0.52 | 0.25 | -0.01 | 0.54 |
| Item 12 | I've been feeling loved | -0.60 | 0.26 | 0.14 | 0.70 |
| Item 13 | I've been interested in new<br>things | -0.49 | 0.24 | 0.18 | 0.41 |
| Item 14 | I've been feeling cheerful | -0.49 | 0.27 | -0.73 | 0.68 |

**Legend:** SE = Standard Error

**Table 4.** Frequency of scoring category responses per item in adults with chronic stroke.

| **Item**<br>**number** | **Item description** | **0**<br>**None**<br>**of the**<br>**time** | **1**<br>**Rarely** | **2**<br>**Some**<br>**of the**<br>**time** | **3**<br>**Often** | **4**<br>**All**<br>**of**<br>**the**<br>**time** |
| --- | --- | --- | --- | --- | --- | --- |
| Item 1 | I've been feeling optimistic about<br>the future | 0 | 2 | 9 | 16 | 10 |
| Item 2 | I've been feeling useful | 0 | 5 | 13 | 15 | 4 |
| Item 3 | I've been feeling relaxed | 0 | 3 | 15 | 13 | 6 |
| Item 4 | I've been feeling interested in other<br>people | 0 | 2 | 7 | 17 | 11 |
| Item 5 | I've had energy to spare | 1 | 7 | 16 | 12 | 1 |
| Item 6 | I've been dealing with problems<br>well | 0 | 1 | 14 | 14 | 8 |
| Item 7 | I've been thinking clearly | 0 | 0 | 10 | 16 | 11 |
| Item 8 | I've been feeling good about<br>myself | 0 | 1 | 10 | 16 | 10 |
| Item 9 | I've been feeling close to other people | 0 | 1 | 9 | 17 | 10 |
| Item 10 | I've been feeling confident | 0 | 2 | 11 | 15 | 9 |
| Item 11 | I've been able to make up my own mind about things | 0 | 1 | 6 | 12 | 18 |
| Item 12 | I've been feeling loved | 0 | 1 | 5 | 12 | 19 |
| Item 13 | I've been interested in new things | 0 | 3 | 6 | 13 | 15 |
| Item 14 | I've been feeling cheerful | 0 | 2 | 7 | 16 | 12 |

**Fig. 3.**
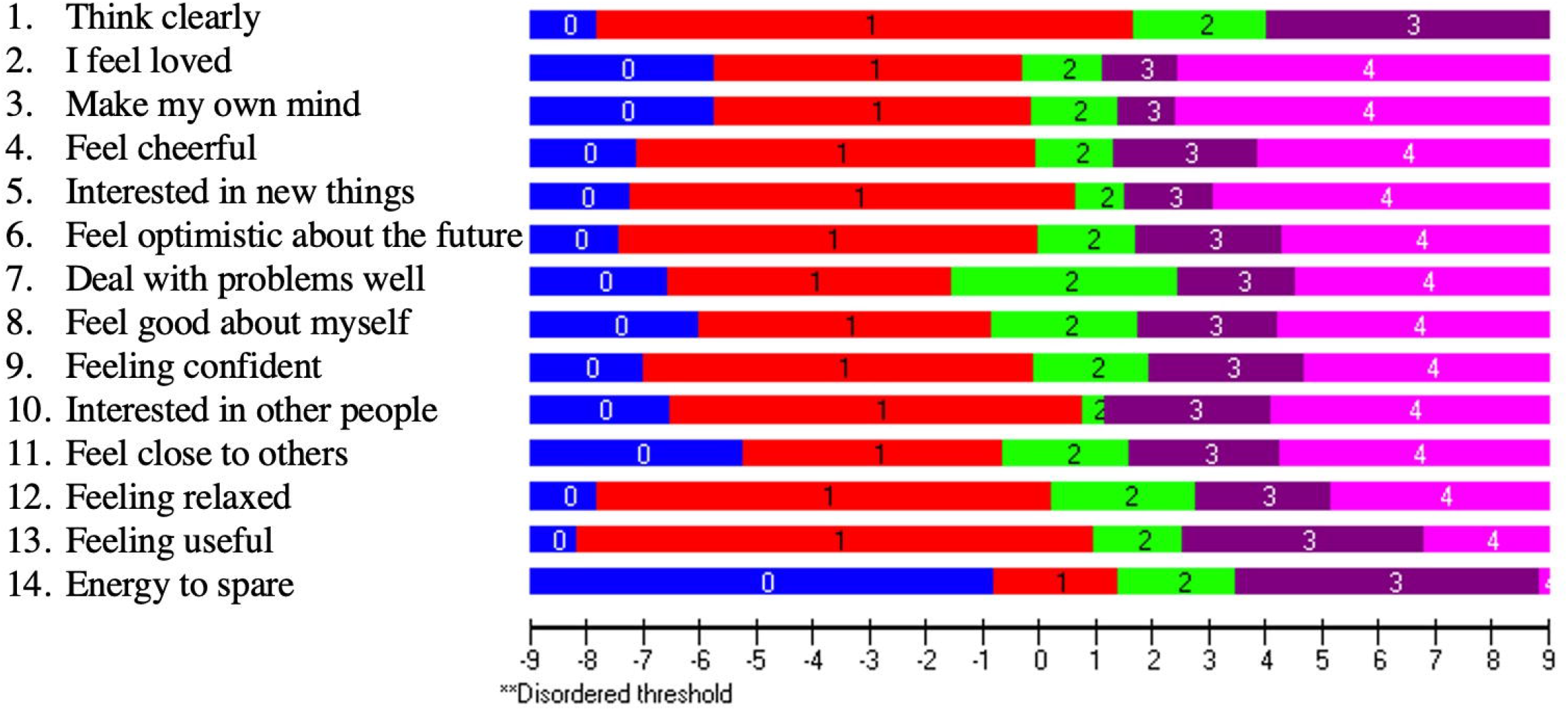
Item threshold map in adults with chronic stroke. The item threshold map shows the hierarchy of the item difficulty levels, with the easiest item on top (item 7 “*I’ve been thinking clearly*”) and the hardest item at the bottom (item 5 “*I’ve had energy to spare*”). The horizontal logit ruler demonstrates the person’s ability level of their positive mental health.

**Fig. 4.**
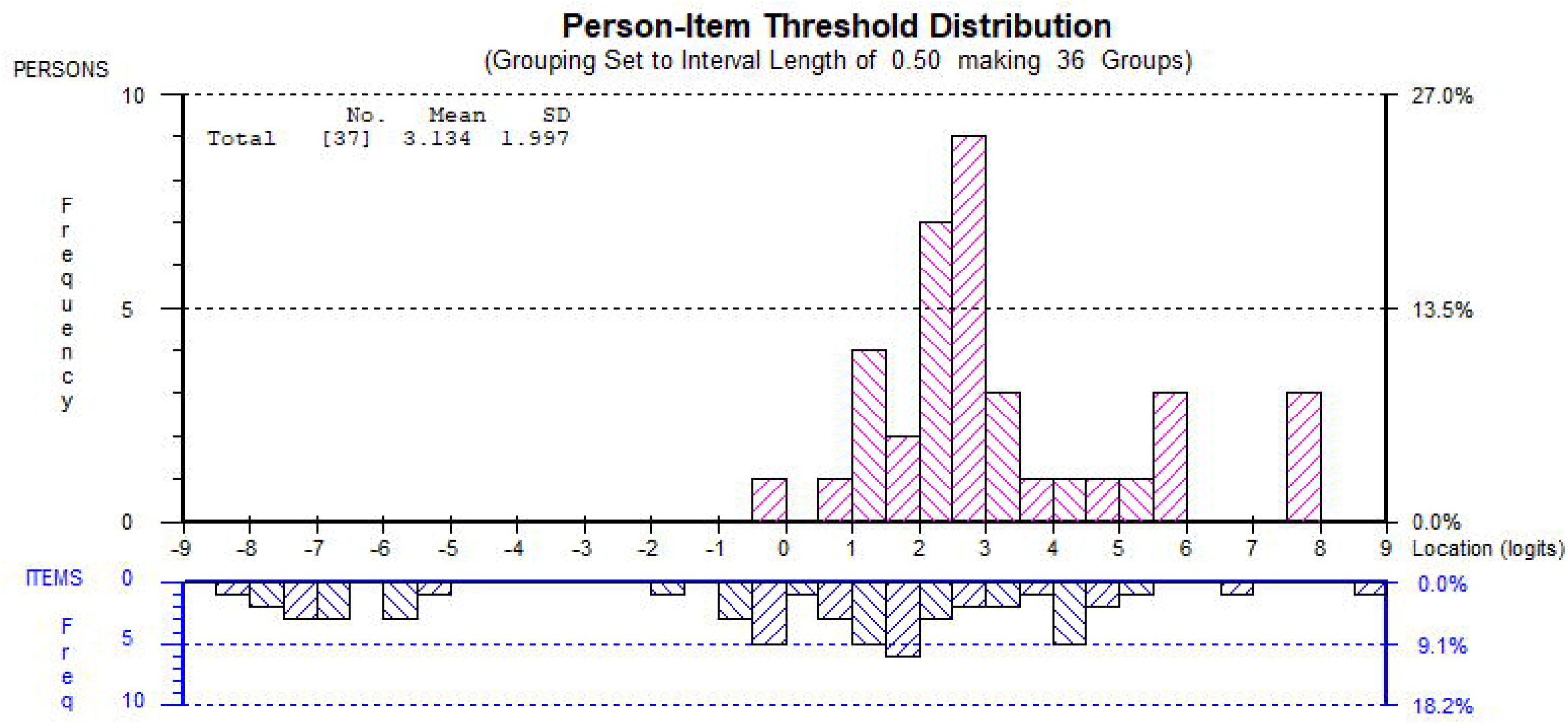
Person-item threshold distribution in adults with chronic stroke. The horizontal logit ruler represents both item difficulty and person ability. The pink histograms show the frequencies of the person’s ability level in terms of positive mental well-being. A higher logit value indicates the person has a higher level of positive mental well-being. The blue histograms represent the frequencies of item difficulty level, and the items are organized from the easiest on the left to the hardest on the right.

Similar to above, we deleted items to match the 7-item SWEMWBS reported in prior studies. After rescoring item 7 to scoring categories [00123], the eigenvalue of the PCAR was 1.79 with 25.51% variance explained by the first component, with paired t-tests showing that 5.41% of pairs had significantly different person locations. This is very close to demonstrating the unidimensionality of the scale. However, the PSR dropped to 0.81, which is not reliable enough for individual decision-making. The person mean location remained high at 3.13±1.84 indicating that the items were too easy. Similar to above, we do not recommend using the 7-item scale. We recommend solving the targeting, as well as validating our results in a larger sample, prior to using the WEMWBS for clinical or research purposes in adults with stroke.

## Discussion

The aims of this study were to investigate the structural validity of the WEMWBS in community-dwelling adults as well as adults with stroke in the US. The WEMWBS shows good item and person fit in both these groups. The main problem was the targeting, demonstrating that the items were too easy for either group. These findings were consistent with the findings in all other studies that reported on person mean locations with Rasch analysis, except for Stewart-Brown et al. (2009), who reported good targeting.

Of note, item fit in the community-dwelling adult group was obtained after deleting misfitting items 1 and 5. Deleting item 5 “*I’ve had energy to spare*” was consistent with earlier studies.[6, 7, 9] In Houghton et al. (2015) item 5 was deleted because DIF was identified for age, while item 5 demonstrated misfit in both Stewart-Brown *et al*. (2009) and Bartram *et al*. (2013). Item 1 “*I have been feeling optimistic about the future*” was maintained in prior studies. During a qualitative study on item comprehension of the WEMWBS, a focus group in Pakistan noticed difficulties in answering “Feeling optimistic about the future”, because there is no translation for “optimistic” in Pashtun.[27] Teenagers in Northern Ireland also expressed difficulty in answering item 1.[28] We did not perform a qualitative analysis after this study and thus were unable to identify the reason for misfit in our US groups. The PCAR analysis pointed to underlying dimensions underneath positive mental health. The items that loaded positively on the first principal component – items 4 “*I have been feeling interested in other people”*, 9 *“I have been feeling close to other people”*, and 12 *“I have been feeling loved”*– all seemed to point to positive feelings regarding interpersonal relationships. The items that loaded negatively on the first principal component seem more related to eudaimonic aspects of life in terms of a person feeling productive regarding their goals and feeling in control of their lives. These were items 6 *“I have been dealing with problems well”*, 7 *“I have been thinking clearly”*, and 8 *“I have been feeling good about myself”*.

In our pilot Rasch analysis in adults with chronic stroke, item fit was obtained after rescoring item 7. The items were too easy for this group as well. Our results in adults with chronic stroke are consistent with Ostir *et al*. (2008), who found that a third of adults reported high levels of positive emotions after 3 months of stroke.[15] The positively loaded items on the first principal component were the same as in the community-dwelling adults. The negatively loaded items were items 1 *“I’ve been feeling optimistic about the future”*, 5 *“I’ve had energy to spare”*, 8 *“I’ve been feeling good about myself”*, and 10 *“I’ve been feeling confident”*. Those items seem to relate to being in control of one’s life after a stroke. It makes sense that the positive feelings about interpersonal relationships might point to something different than having control over one’s life.

Even though studies evaluating the WEMWBS were conducted in different countries with inherent differences in culture, it is noteworthy that across several studies, including our own studies, items 11 *“I’ve been able to make up my own mind about things”* and 7 *“I have been thinking clearly”* were consistently rated as the easiest items, whereas items 5 “*I’ve had energy to spare*” and 3 *“I’ve been feeling relaxed”* were consistently rated as the hardest items.[7, 9]

## Conclusions

The WEMWBS demonstrated good item fit and person fit in a US population of community-dwelling adults and adults with stroke. However, the items are too easy, which is a consistent finding across the majority of WEMWBS Rasch studies performed in different countries. Thus, including more difficult items in a next iteration of the scale could help solve the targeting. Finally, further studies with larger sample sizes should be performed in adults with stroke, and perhaps include adults with acute and subacute stroke, to validate our preliminary findings.

## Data Availability

The data are deposited on the Data Repository for the U of Minnesota (DRUM).

https://doi.org/10.13020/jdfb-pn26

## List of abbreviations

WEMWBS: Warwick-Edinburgh Mental Well-being Scale
SWEMWBS: Short Warwick Edinburgh Mental Well-Being Scale
RUMM: Rasch Unidimensional Measurement Model
UBACC: University of California, San Diego Brief Assessment of Capacity to Consent
PSR: Person separation reliability
DIF: Differential item functioning
PCAR: Principal components analysis of residuals

## Declaration

### Ethics and approval and consent to participate

The study was approved by the University of Minnesota’s Institutional Review Board (IRB# STUDY00005849 and STUDY00000821) and the study was in accordance with the Declaration of Helsinki. All of the healthy adults agreed with study consent and they were quizzed on the comprehension of the content of the consent form through the University of California, San Diego Brief Assessment of Capacity to Consent (UBACC).

### Consent for publication

N/A

### Availability of data and materials

The dataset(s) supporting the conclusions of this article is(are) available in the Data Repository for U of M (DRUM), https://doi.org/10.13020/jdfb-pn26

### Authors’ contributions

All authors contributed substantially to parts of the manuscript, and critically revised it for content, approved the final version, and agreed to be accountable for the accuracy and integrity of this work. Specific contributions include:

- Conception or design of the work: AVDW
- Acquisition and analysis of evidence: AVDW, WD
- Interpretation of the evidence: AVDW, WD, SC, JB

## Acknowledgements

We appreciate all the participants who have participated in the study, and the research volunteers who have helped with data collection. Our profound gratitude goes to Marc Noël for the critical review of the manuscript.

## STROBE Statement—Checklist of items that should be included in reports of *cross-sectional studies*

| PRoBES Statement – Checklist of items that should be included in reports of cross-sectional studies |  |  | Page No |
| --- | --- | --- | --- |
|  | Item No | Recommendation |  |
| Title and abstract | 1 | (a) Indicate the study’s design with a commonly used term in the title or the abstract | 1 |
|  |  | (b) Provide in the abstract an informative and balanced summary of what was done and what was found | 2-3 |
| Introduction |  |  |  |
| Background/rationale | 2 | Explain the scientific background and rationale for the investigation being reported | 4-6 |
| Objectives | 3 | State specific objectives, including any prespecified hypotheses | 6 |
| Methods |  |  |  |
| Study design | 4 | Present key elements of study design early in the paper | 6 |
| Setting | 5 | Describe the setting, locations, and relevant dates, including periods of recruitment, exposure, follow-up, and data collection | 6 |
| Participants | 6 | (a) Give the eligibility criteria, and the sources and methods of selection of participants | 6-7 |
| Variables | 7 | Clearly define all outcomes, exposures, predictors, potential confounders, and effect modifiers. Give diagnostic criteria, if applicable | 7 |
| Data sources/ measurement | 8* | For each variable of interest, give sources of data and details of methods of assessment (measurement). Describe comparability of assessment methods if there is more than one group | 7 |
| Bias | 9 | Describe any efforts to address potential sources of bias | 7 |
| Study size | 10 | Explain how the study size was arrived at | N/A |
| Quantitative variables | 11 | Explain how quantitative variables were handled in the analyses. If applicable, describe which groupings were chosen and why | N/A |
| Statistical methods | 12 | (a) Describe all statistical methods, including those used to control for confounding | 7-8 |
|  |  | (b) Describe any methods used to examine subgroups and interactions | N/A |
|  |  | (c) Explain how missing data were addressed | N/A |
|  |  | (d) If applicable, describe analytical methods taking account of sampling strategy | N/A |
|  |  | (e) Describe any sensitivity analyses | N/A |
| Results |  |  |  |
| Participants | 13* | (a) Report numbers of individuals at each stage of study—eg numbers potentially eligible, examined for eligibility, confirmed eligible, included in the study, completing follow-up, and analysed | N/A |
|  |  | (b) Give reasons for non-participation at each stage | N/A |
|  |  | (c) Consider use of a flow diagram | N/A |
| Descriptive data | 14* | (a) Give characteristics of study participants (eg demographic, clinical, social) and information on exposures and potential confounders | 9 |
|  |  | (b) Indicate number of participants with missing data for each variable of interest | N/A |
| Outcome data | 15* | Report numbers of outcome events or summary measures | Table 2.<br>Table 3.<br>Table 4.<br>Additional<br>file. |
| Main results | 16 | (a) Give unadjusted estimates and, if applicable, confounder-adjusted estimates and their precision (eg, 95% confidence interval). Make clear which confounders were adjusted for and why they were included | N/A |
|  |  | (b) Report category boundaries when continuous variables were categorized | N/A |
|  |  | (c) If relevant, consider translating estimates of relative risk into absolute risk for a meaningful time period | N/A |
| Other analyses | 17 | Report other analyses done—eg analyses of subgroups and interactions, and sensitivity analyses | N/A |
| **Discussion** |  |  |  |
| Key results | 18 | Summarise key results with reference to study objectives | 12 |
| Limitations | 19 | Discuss limitations of the study, taking into account sources of potential bias or imprecision. Discuss both direction and magnitude of any potential bias | 14 |
| Interpretation | 20 | Give a cautious overall interpretation of results considering objectives, limitations, multiplicity of analyses, results from similar studies, and other relevant evidence | 12-14 |
| Generalisability | 21 | Discuss the generalisability (external validity) of the study results | 14 |
| **Other information** |  |  |  |
| Funding | 22 | Give the source of funding and the role of the funders for the present study and, if applicable, for the original study on which the present article is based | 16 |

\*Give information separately for exposed and unexposed groups.
**Note:** An Explanation and Elaboration article discusses each checklist item and gives methodological background and published examples of transparent reporting. The STROBE checklist is best used in conjunction with this article (freely available on the Web sites of PLoS Medicine at http://www.plosmedicine.org/, Annals of Internal Medicine at http://www.annals.org/, and Epidemiology at http://www.epidem.com/). Information on the STROBE Initiative is available at www.strobe-statement.org.

